# Time to PrEP Disengagement and Associated Factors Among Adolescents and Young Adults from Key and Priority Populations in Uganda. A survival Analysis

**DOI:** 10.64898/2026.07.22.26358724

**Authors:** Simon Mwima, Stephen Walwo

## Abstract

**Background:** Adolescents and young adults (AYAs) from key and priority populations face persistent challenges with sustained engagement in HIV pre-exposure prophylaxis (PrEP) care. While PrEP initiation has expanded across sub-Saharan Africa, evidence on long-term retention and determinants of disengagement among AYAs remains limited. We examined time to PrEP disengagement and associated factors among AYAs initiating PrEP in eastern Uganda.

**Methods:** We conducted a retrospective longitudinal analysis of routinely collected program data for AYAs aged 15–29 years from key and priority populations who initiated PrEP between 2019 and 2025 at Mbale Regional Referral Hospital. Time to PrEP disengagement was assessed using Kaplan– Meier survival analysis and Cox proportional hazards regression. Multivariable models adjusted for sociodemographic, relational, behavioral, and service delivery factors. Sensitivity analyses redefined the time origin to day 91 following PrEP initiation to reflect the programmatic 90-day grace period.

**Results:** Among 3,553 AYAs initiating PrEP, the median time to disengagement was 284 days (95% CI: 273–295). The median age was 24 years (interquartile range [IQR]: 20–26). The probability of remaining engaged in PrEP care declined from 60.1% at 90 days to 20.2% at 365 days. Survival patterns differed significantly by population category and sex at birth but not by age group. In adjusted analyses (N = 3,391), knowledge of a partner’s HIV status (aHR = 2.04; 95% CI: 1.82– 2.29) and initiation through community-based services (aHR = 1.42; 95% CI: 1.17–1.72) were associated with faster disengagement. Married participants had lower hazards of disengagement compared with single participants (aHR = 0.69; 95% CI: 0.64–0.76). Reporting an STI syndrome (aHR = 0.42; 95% CI: 0.32–0.55) or recent gender-based violence (aHR = 0.76; 95% CI: 0.60– 0.96) was associated with reduced disengagement. Findings were highly consistent in sensitivity analyses using an alternative risk-period definition.

**Conclusions:** PrEP disengagement among AYAs occurs rapidly following initiation, with substantial attrition within the first year. Relational factors, service delivery modality, and population-specific vulnerabilities strongly shape retention trajectories. These findings underscore the need for risk-responsive, differentiated PrEP delivery strategies that strengthen partner-based services, integrate STI and GBV screening, and adapt retention support for AYAs in community and facility settings.

## Background

Oral pre-exposure prophylaxis (PrEP) is a highly efficacious biomedical intervention for preventing HIV acquisition (Corneli et al., 2014). Its effectiveness has been demonstrated across multiple randomized controlled trials and open-label studies among populations at substantial risk of HIV infection (Corneli et al., 2014; Grant et al., 2010). When taken consistently, daily oral PrEP reduces HIV acquisition by more than 90%, underscoring its potential to substantially curb HIV incidence in high-burden settings (McCormack et al., 2016). Studies among populations at high risk of HIV in sub-Saharan Africa, including sex workers, men who have sex with men, fisherfolk, and HIV-discordant couples, have reported high initiation rates, ranging from 60% to 90% (Baeten et al., 2016; Heffron et al., 2012). However, subsequent PrEP demonstration projects have reported low retention, with follow-up rates as low as approximately 14–40% in some African settings and 28–37% by 3–6 months post-initiation, highlighting challenges in sustaining engagement outside controlled trial environments (Cowan et al., 2018; Eakle et al., 2017; Kyongo et al., 2018).

In Uganda, while there has been expanded availability and programmatic scale-up of PrEP, substantial losses persist at later stages of the PrEP continuum (Kagaayi et al., 2020; UNAIDS, 2025). By the end of 2023, the country had recorded approximately 550,000 individuals who had initiated oral PrEP; however, sustained engagement remains low. A national gap analysis reported that fewer than 30% of the cumulative number of PrEP users were refilled each quarter (Rosen et al., 2025). Early disengagement from PrEP care is common and undermines the population-level effectiveness of PrEP. Disengagement may occur due to dynamic HIV risk perception, changes in sexual partnerships, reluctance or difficulty maintaining daily medication use, stigma, and competing social and economic priorities (Mwima et al., 2025; Zimmermann et al., 2019). Adolescents and young adults may be particularly vulnerable to disengagement given high mobility, evolving risk profiles, and work-related transitions, and heightened exposure to stigma within both community and facility-based settings (Kawuma et al., 2025).

Despite these vulnerabilities, longitudinal patterns of PrEP disengagement remain poorly described in Uganda, especially among young people from key populations (Kagaayi et al., 2020). Existing evidence has largely relied on short follow-up periods, cross-sectional designs, or narrowly defined cohorts, limiting the ability to characterize the timing and trajectories of disengagement over extended periods (Kagaayi et al., 2020; Kawuma et al., 2025; Ntabadde et al., 2024). Programmatic data from early PrEP rollout among key and priority populations demonstrated that disengagement occurred rapidly after initiation, with a median time on PrEP of approximately 45 days, indicating that half of initiators disengaged within the first two months of care (Kagaayi et al., 2020). More recent facility-based evidence among female sex workers similarly showed substantial attrition, with fewer than half of participants retained in care by six months following PrEP initiation (Kawuma et al., 2025). Together, these studies indicate that PrEP disengagement is common and often occurs early in Uganda; however, limitations related to short follow-up, single-population or single-site designs, and limited longitudinal measurement persist. Consequently, little is known about long-term time to PrEP disengagement among adolescents and young adults from key populations, or how programmatically actionable individual and service-level factors shape disengagement risk across differentiated service delivery platforms in routine care settings.

In the current study, we address this critical implementation gap by examining time to PrEP disengagement among adolescents and young adults accessing PrEP through facility and community-based service delivery models in eastern Uganda over a five-year period. Using routinely collected patient data and time-to-event methods, we characterize longitudinal patterns of engagement and identify factors associated with early PrEP discontinuation. By focusing on sustained engagement in real-world program settings, this study provides policy-relevant evidence to inform differentiated, risk-responsive retention strategies and strengthen the effectiveness of PrEP programs beyond initiation among populations at highest risk of HIV infection.

## Conceptual framework

This study was guided by an integrated conceptual framework combining the Modified Social Ecological Model (Baral et al., 2013) and the PrEP Continuum Cascade (Nunn et al., 2017). The PrEP cascade conceptualizes engagement as a sequence from initiation through persistence to discontinuation, highlighting that the preventive impact of PrEP depends on sustained use over time rather than on initiation alone. Accordingly, this analysis focuses on the persistence stage of the cascade, operationalized as time to PrEP disengagement following initiation. MSEM situates PrEP persistence within a multilevel context, recognizing that disengagement is shaped by interacting individual, interpersonal, and structural factors. Guided by this framework, covariates were grouped into sociodemographic factors (e.g., age, sex at birth, education level, marital status); behavioral and vulnerability-related factors (e.g., partner HIV testing status, exposure to gender-based violence) and structural and programmatic factors, including key population classification (e.g., sex worker, MSM, PWID) and service delivery model (facility and community). Integrating SEM with the PrEP continuum framework provides a theory-driven approach to examining PrEP disengagement under routine implementation conditions among adolescents and young adults from key and priority populations.

## Methods and materials

### Study design and setting

We conducted a retrospective longitudinal cohort analysis using routinely collected program data from HIV prevention services at Mbale Regional Referral Hospital (MRRH) in eastern Uganda. Mbale region is characterized by substantial HIV vulnerability among AYAs, particularly those from key and priority populations (KPs/PPs), including sex workers, men who have sex with men, people who inject drugs, and other young people engaged in high-risk sexual behaviors. These populations experience a disproportionate burden of HIV risk due to intersecting behavioral, social, and structural factors such as stigma, mobility, limited access to youth-friendly services, and economic vulnerability (UNAIDS, 2023; MoH, 2022). In response to these vulnerabilities, MRRH hosts a fully established adolescent and young people–focused HIV clinic that also functions as a drop-in center (DIC) providing stigma-free, youth-friendly, and KP-responsive HIV prevention services.

### The PrEP programme

In 2019, the Ministry of Health Uganda expanded access to oral pre-exposure prophylaxis, tenofovir disoproxil fumarate (TDF) combined with lamivudine (3TC), to include adolescents and young people at substantial risk of HIV infection (MoH, 2022a, 2022b). The programme is implemented through selected PrEP-accredited sites, including Mbale Regional Referral Hospital in eastern Uganda, with financial and technical support from the government of Uganda, the Global Fund to Fight AIDS, Tuberculosis and Malaria and PEPFAR through the Centers for Disease Control and Prevention (MoH, 2022a). PrEP Eligibility is determined using a Ministry of Health, approved HIV risk screening tool, with individuals classified as being at substantial risk if they report at least one high-risk sexual or drug-use exposure (MoH, 2022b). PrEP services are delivered through a differentiated model that combines facility and community-based approaches, consistent with national guidance on key population programming. Facility-based services are provided at the hospital HIV clinic, while community-based delivery includes mobile outreaches and hotspot-based services supported by trained peer educators. To minimize stigma, community mobilization emphasizes PrEP availability for individuals at substantial HIV risk without explicitly naming population categories, relying on peer-led, network-based approaches consistent with chain-referral principles. At both facility and outreach sites, multidisciplinary teams provide HIV testing, risk assessment, and clinical screening in accordance with national guidelines, including STI assessment and renal function screening. Client follow-up includes a mandatory 30-day refill and reassessment visit, with subsequent visits scheduled flexibly based on client preferences, drug availability, and clinical considerations. Follow-up services include PrEP refills, adherence counselling, HIV retesting, and ongoing risk reassessment. Programme data, including sociodemographic characteristics, service delivery modality, key and priority population classification, STI assessment, partner HIV status knowledge, and exposure to gender-based violence, are routinely captured in KP tracker and electronic HMIS records. The present study represents a secondary analysis of these routinely collected programmatic data.

### Inclusion and exclusion criteria

The analytic sample comprised AYAs aged 15–29 years from key and priority populations who accessed HIV prevention services between 2019 and 2025 at Mbale Regional Referral Hospital and initiated PrEP. Individuals were included if they had a valid PrEP initiation date and sufficient follow-up information to establish time to disengagement or censoring. Participants were excluded if they had missing or invalid time variables (including negative follow-up intervals) or incomplete baseline records that precluded determination of time-to-event status. Clients who re-initiated PrEP after a documented disengagement episode were retained in the dataset; however, time-to-disengagement was defined from the first PrEP initiation episode, and subsequent re-initiations were not treated as separate risk intervals. Participants who remained active on PrEP at the end of follow-up, transferred out of services, had a future scheduled return date, or were otherwise not observed to disengage were right-censored at their last recorded visit, applying a 90-day grace period.

### Data source and quality

This analysis used routinely collected secondary data from the Ministry of Health–approved national KP tracker. Data is entered during routine service delivery, with system-embedded validation and mandatory fields to support data quality. Extracted datasets were checked for completeness, internal consistency, and duplicates, in accordance with Ministry of Health quality assurance procedures (MoH, 2020; KP Tracker).

### Data management and missing data

Extracted data was cleaned and prepared for analysis using standardized coding and verification procedures. Unique client identifiers were used to confirm record linkage across visits and to prevent duplication. Missing data were assessed descriptively by variable and outcome status. Key demographic variables had minimal missingness due to system-enforced mandatory fields within the national KP tracker. Analyses were conducted using complete cases for model-specific variables. No imputation was performed, as the data were routinely collected for programmatic purposes and missingness primarily reflected routine service documentation rather than study-related data collection.

### Analytic sample and censoring

The analytic sample included 3,553 individuals who initiated PrEP during the study period and had a documented baseline visit in the national KP tracker. Disengagement from PrEP was defined as a 90-day grace period from the last documented visit. During follow-up, 3,401 (95.7%) individuals met the criteria for disengagement, while 152 (4.3%) remained active and were censored. Participants were right censored at the earliest of the following: the last documented PrEP follow-up visit without evidence of disengagement, a future return date, a documented transfer out of PrEP services, or administrative censoring at the end of the study period (December 2, 2025). Individuals who remained actively engaged in PrEP care at the end of follow-up were treated as censored observations.

### Outcome variable

The primary outcome was time to PrEP disengagement, defined as the number of days from PrEP initiation to disengagement from PrEP care (Lichtwarck et al., 2023). Disengagement was operationalized using a return date-based definition with a 90-day grace period, whereby individuals were classified as disengaged if they failed to return within 90 days beyond their expected return date recorded at the most recent PrEP visit. Event status was binary-coded (1 = disengaged from PrEP; 0 = right-censored).

### Independent variable and covariates

Primary explanatory variables included service delivery model (0 = facility-based and 1 = community-based) and population category, classified according to national key and priority population definitions and modeled as categorical variables with sex workers as the reference group (men who have sex with men, people who inject drugs, and other priority populations entered as indicator variables). Sociodemographic covariates included age group (15–19, 20–24, 25–29 years), sex at birth (female, male), education level with primary as the reference group (primary, secondary, tertiary), marital status with singles as the reference group (single, married, previously married), and religion (Christian, Muslim). Baseline vulnerability and service-related indicators included knowledge of partner’s HIV status coded as 0 = does not know and 1 = knows partner’s HIV status, peer follow-up after PrEP initiation, coded as 0 = no and 1 = yes, sexually transmitted infection screening status (positive vs negative), reported psychoactive drug use (yes vs no), and exposure to gender-based violence (yes vs no). All variables were measured at initiation, selected a priori based on theory, existing evidence, and programmatic relevance, and included in multivariable models to adjust for potential confounding.

### Ethical considerations

This study was a secondary analysis of de-identified routine program data. Authorization was obtained from the Uganda Ministry of Health (Ref: ADM/130/313/05), and ethical approval was granted by the Mbale Regional Referral Hospital Research Ethics Committee (MRRH-2025-552) in February 2025. A waiver of informed consent was approved in accordance with ethical guidelines. Data were securely stored and accessed only by authorized personnel, and the study was conducted in line with the Declaration of Helsinki and applicable Ugandan regulations.

## Results

### Baseline descriptive characteristics

A total of 3,553 adolescents and young adults aged 15–29 years from key and priority populations initiated PrEP during the study period (Table 1). Slightly over half of participants were female (53.1%, n = 1,888), while (46.9%, n = 1,665) were male. More than half (53.3%, n = 1,895) had attained secondary education; (40.8%, n = 1,451) had none or primary education; and a small proportion had tertiary education (5.8%, n = 207). Majority of participants identified as Christian (73.7%, n = 2,557), while (26.3%, n = 911) were Muslim. Nearly half of the cohort was single (47.7%, n = 1,694), (36.2%, n = 1,286) were married, and (16.1%, n = 573) were previously married (divorced, separated, or widowed). In terms of population classification, sex workers constituted the largest group (50.1%, n = 1,779), followed by men who have sex with men (34.6%, n = 1,229), people who inject drugs (7.9%, n = 279), and priority populations (7.5%, n = 266). Young adults aged 20–24 years represented the largest age group (42.6%, n = 1,514), followed by those aged 25–29 years (40.5%, n = 1,439), while adolescents aged 15–19 years accounted for (16.9%, n = 600) of participants. With respect to relationship and vulnerability characteristics, the majority of participants did not know their partner’s HIV status (86.4%, n = 3,071), while (13.6%, n = 482) reported knowing their partner’s status. Most participants had been contacted by a peer outreach worker (83.1%, n = 2,953). Reported gender-based violence (GBV) was uncommon (2.4%, n = 85), and STI syndromes at baseline were identified in (1.7%, n = 61) of those assessed. Any psychoactive drug use in the past six months was reported by (7.1%, n = 251) of participants. The vast majority of PrEP initiations occurred through community-based service delivery points (93.9%, n = 3,335), with only 6.1% (n = 218) initiating PrEP at health facilities.

**Table 1.**
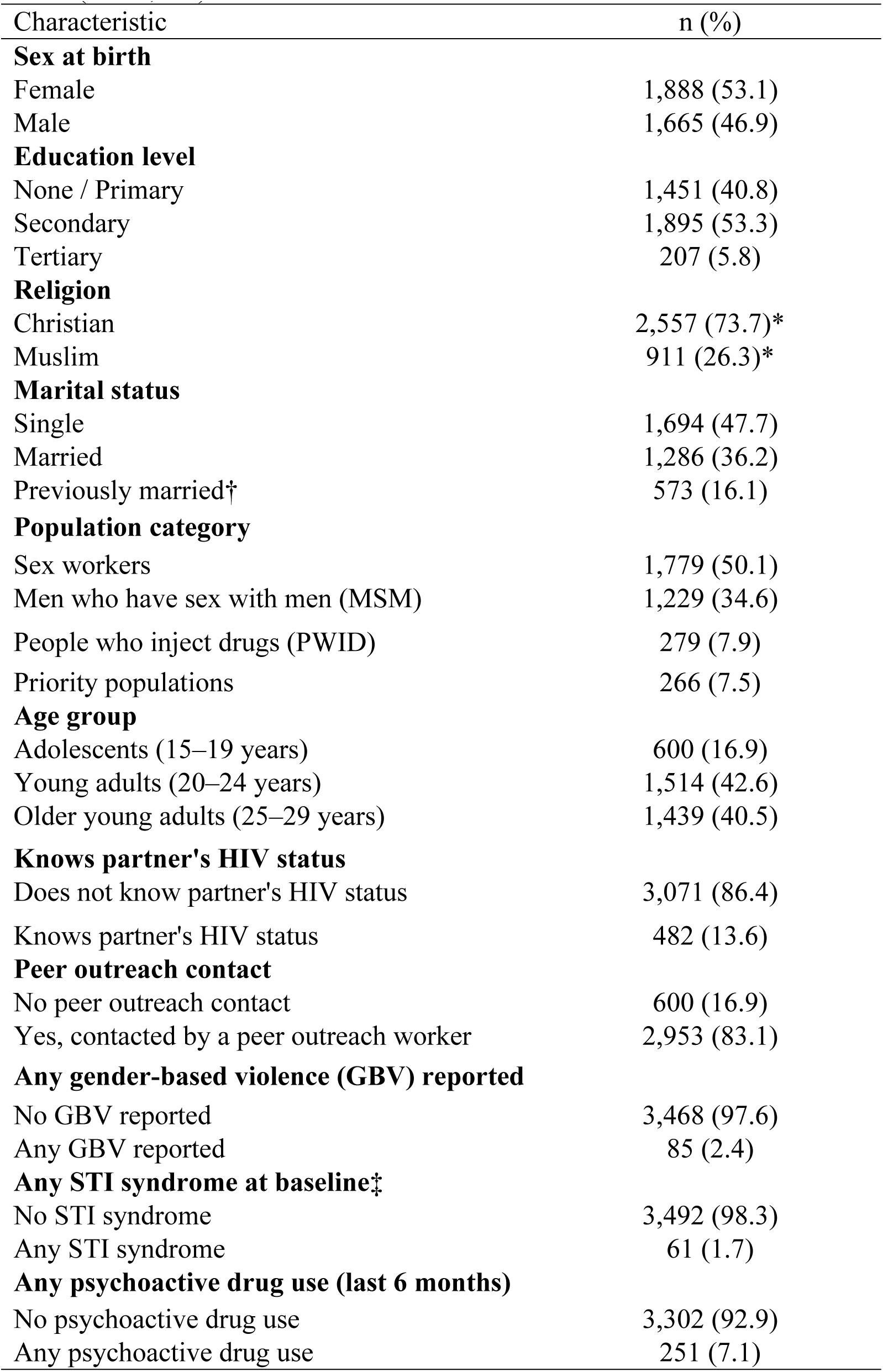
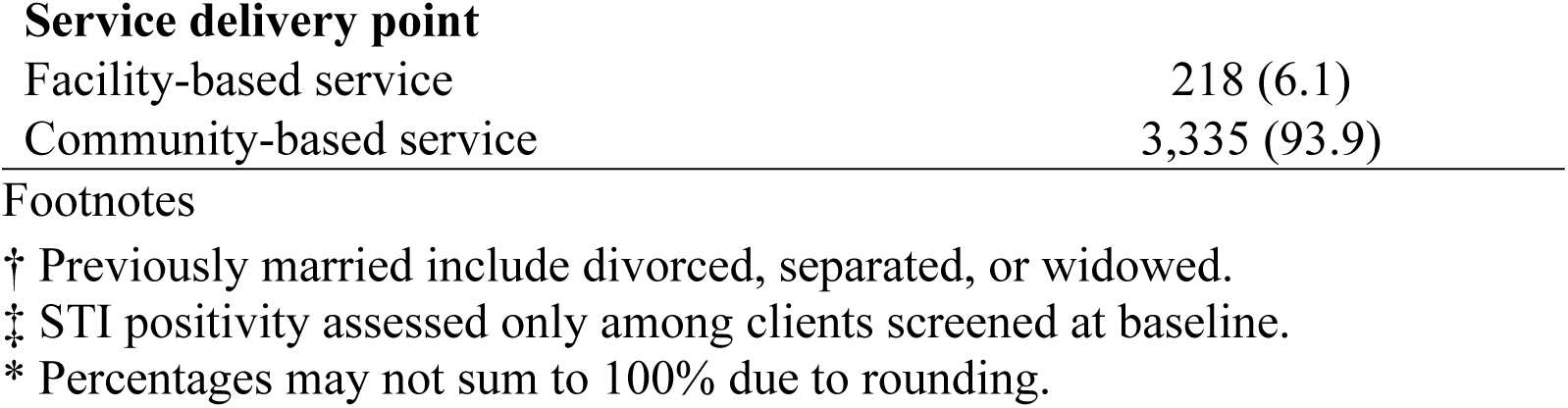
Baseline Characteristics of Adolescents and Young Adults Initiating PrEP (N = 3,553)

| Characteristic | n (%) |
| --- | --- |
| **Sex at birth** |  |
| Female | 1,888 (53.1) |
| Male | 1,665 (46.9) |
| **Education level** |  |
| None / Primary | 1,451 (40.8) |
| Secondary | 1,895 (53.3) |
| Tertiary | 207 (5.8) |
| **Religion** |  |
| Christian | 2,557 (73.7)* |
| Muslim | 911 (26.3)* |
| **Marital status** |  |
| Single | 1,694 (47.7) |
| Married | 1,286 (36.2) |
| Previously married† | 573 (16.1) |
| **Population category** |  |
| Sex workers | 1,779 (50.1) |
| Men who have sex with men (MSM) | 1,229 (34.6) |
| People who inject drugs (PWID) | 279 (7.9) |
| Priority populations | 266 (7.5) |
| **Age group** |  |
| Adolescents (15–19 years) | 600 (16.9) |
| Young adults (20–24 years) | 1,514 (42.6) |
| Older young adults (25–29 years) | 1,439 (40.5) |
| **Knows partner's HIV status** |  |
| Does not know partner's HIV status | 3,071 (86.4) |
| Knows partner's HIV status | 482 (13.6) |
| **Peer outreach contact** |  |
| No peer outreach contact | 600 (16.9) |
| Yes, contacted by a peer outreach worker | 2,953 (83.1) |
| **Any gender-based violence (GBV) reported** |  |
| No GBV reported | 3,468 (97.6) |
| Any GBV reported | 85 (2.4) |
| **Any STI syndrome at baseline‡** |  |
| No STI syndrome | 3,492 (98.3) |
| Any STI syndrome | 61 (1.7) |
| **Any psychoactive drug use (last 6 months)** |  |
| No psychoactive drug use | 3,302 (92.9) |
| Any psychoactive drug use | 251 (7.1) |
| **Service delivery point** |  |
| Facility-based service | 218 (6.1) |
| Community-based service | 3,335 (93.9) |

### Kaplan–Meier survival Analysis of time to PrEP disengagement

The Kaplan–Meier survival curve illustrating overall time to PrEP disengagement is presented in Figure 1. The probability of remaining engaged in PrEP care was 100% at 30 days, 60.1% at 90 days, 45.2% at 180 days, and 20.2% at 365 days following PrEP initiation (Table 2). The median time to PrEP disengagement was 284 days (95% CI: 273–295 days), indicating that half of participants disengaged from PrEP care within approximately nine months of initiation. Survival patterns were broadly similar across age groups (Figure 2a), with Kaplan–Meier curves largely overlapping; supported by the log-rank test, which showed no statistically significant differences in survival distributions by age group (χ² = 2.45, df = 2, p = 0.294). In contrast, Kaplan–Meier curves differed markedly by population category (Figure 2b), with priority populations demonstrating the highest retention over time, followed by MSM and PWID, while sex workers experienced earlier disengagement; these differences were statistically significant (log-rank χ² = 197.36, df = 3, p < 0.001). Survival distributions also differed significantly by sex at birth (Figure 2c), with males exhibiting longer retention than females (log-rank χ² = 35.86, df = 1, p < 0.001). These unadjusted survival patterns were subsequently examined using Cox proportional hazards regression models.

**Figure 1.**
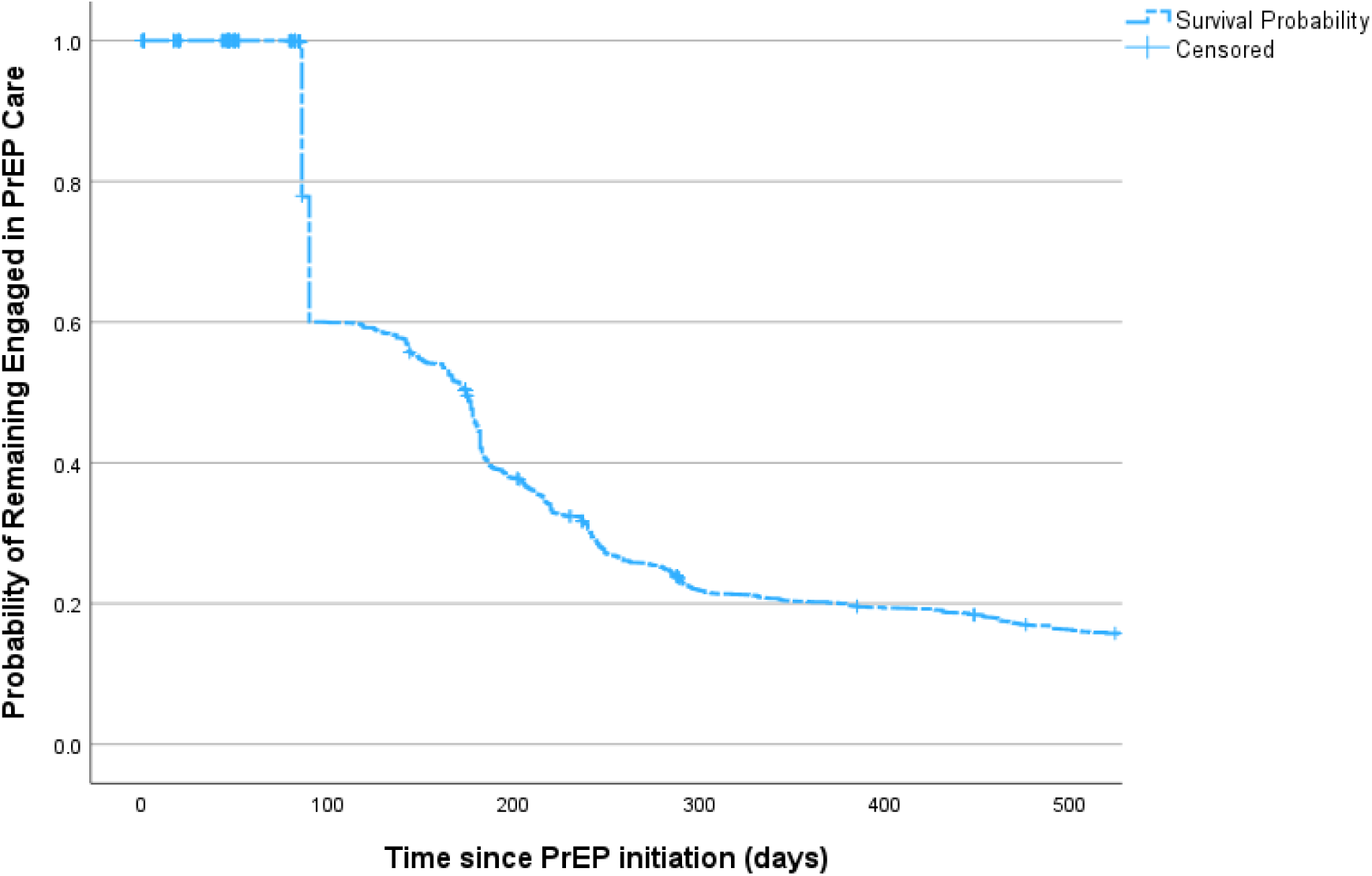
Overall Kaplan–Meier survival estimate of time to PrEP disengagement among key and priority populations. Kaplan–Meier survival curve shows the probability of remaining engaged in PrEP care following initiation. Survival probabilities were 100% at 30 days, 60.1% at 90 days, 45.2% at 180 days, and 20.2% at 365 days. The median time to PrEP disengagement was 284 days (95% CI: 273– 295 days). Tick marks indicate censored observations.

**Table 2.** Survival probabilities for time to PrEP disengagement (N = 3,553)

| Time since PrEP initiation | Survival probability | Standard error |
| --- | --- | --- |
| 30 days | 1 | 0 |
| 90 days | 0.601 | 0.008 |
| 180 days | 0.452 | 0.008 |
| 365 days | 0.202 | 0.007 |
| Median time to disengagement (days) | 284 | 5.48 |

Survival probabilities were estimated using Kaplan–Meier methods. Estimates at fixed time points represent the cumulative probability of remaining engaged in PrEP care beyond the specified time.

**Figure 2a.**
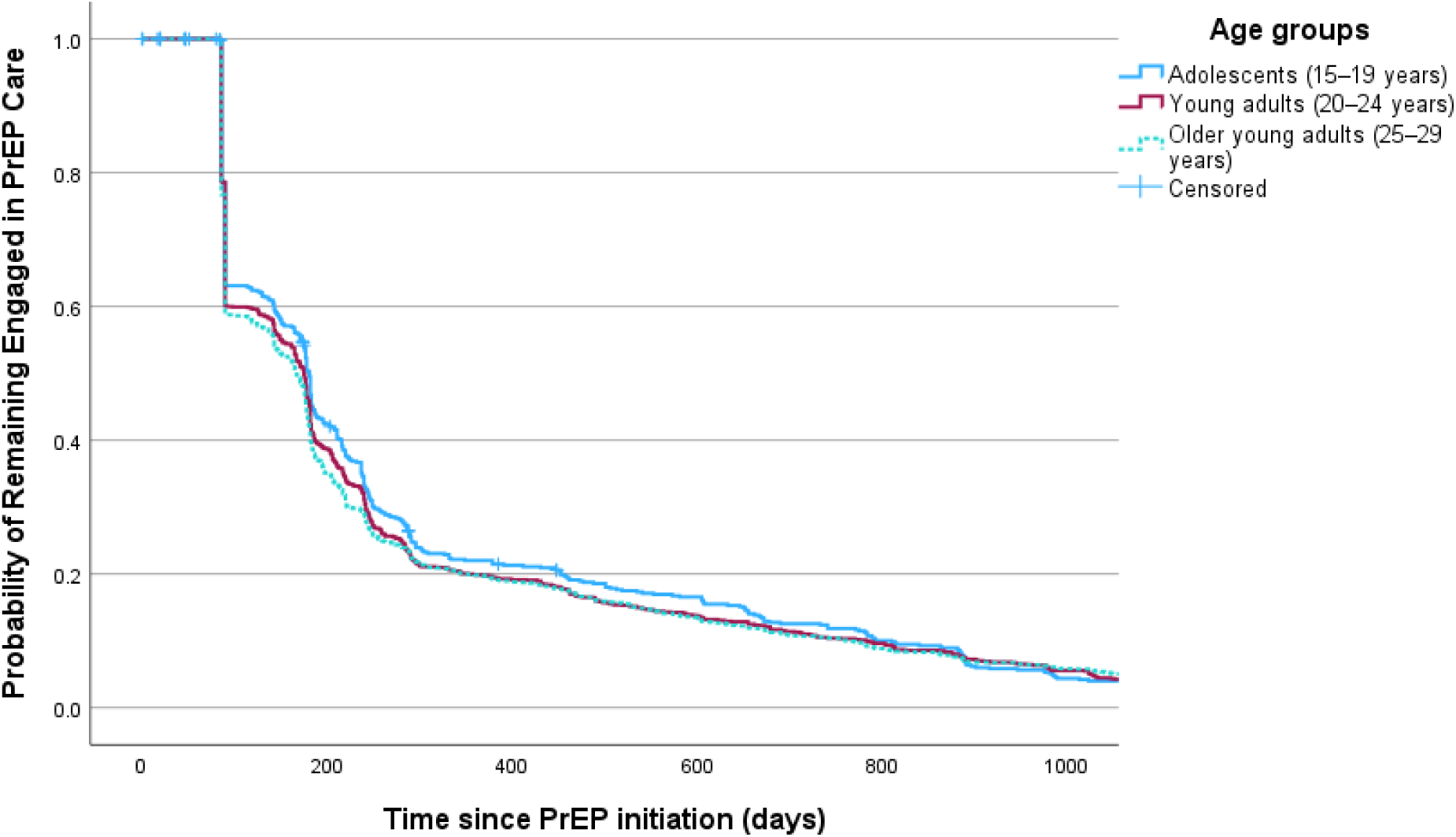
Kaplan–Meier survival curves for time to PrEP disengagement by age group. Kaplan–Meier curves largely overlapping; supported by the log-rank test with no statistically significant differences in survival distributions (χ² = 2.45, df = 2, p = 0.294)

**Figure 2b.**
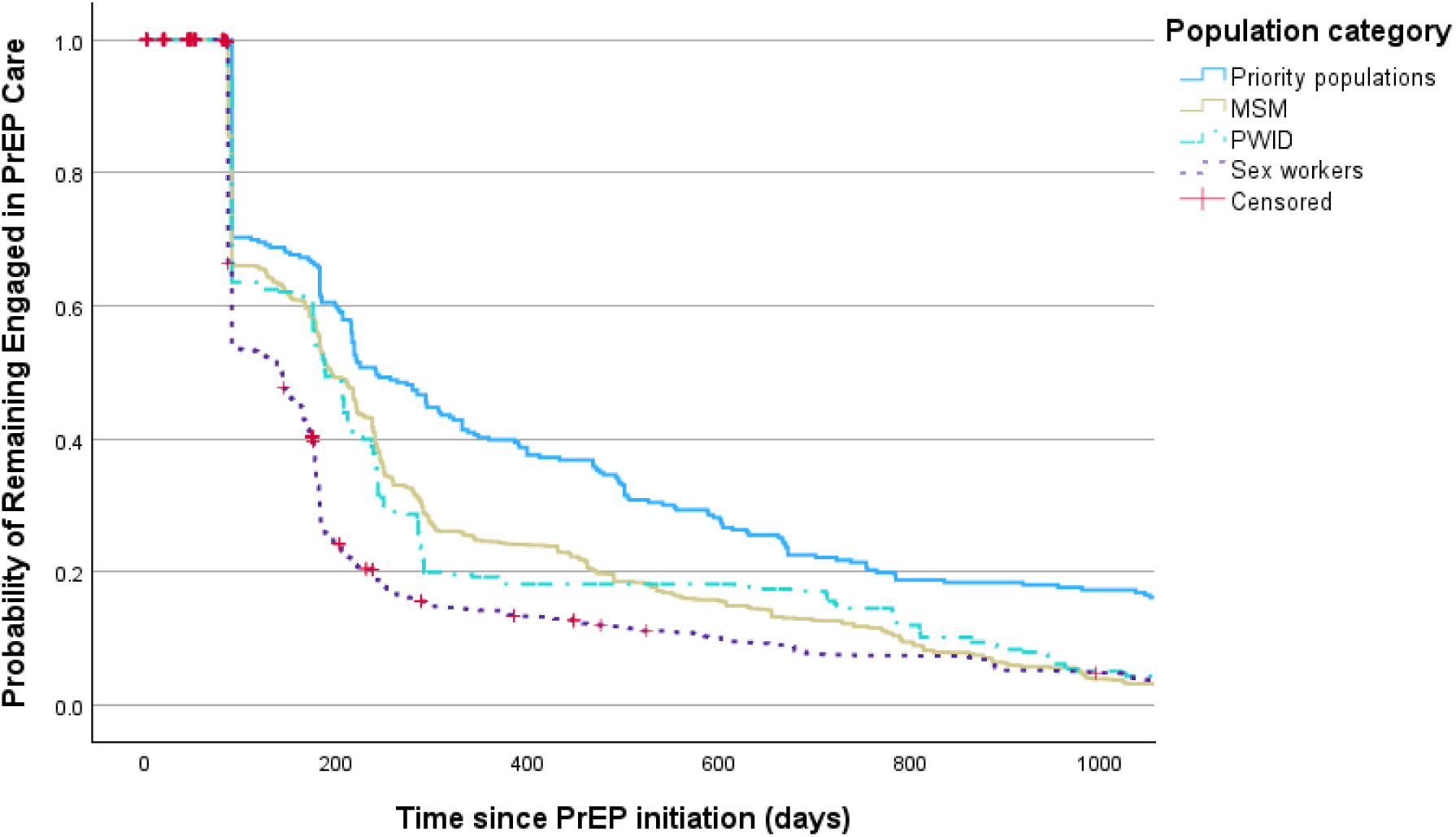
Kaplan–Meier survival curves of time to PrEP disengagement by population category. Kaplan–Meier survival curves differed significantly across population groups (log-rank test, χ² = 197.36, df = 3, p < 0.001). Tick marks indicate censored observations.

**Figure 2c.**
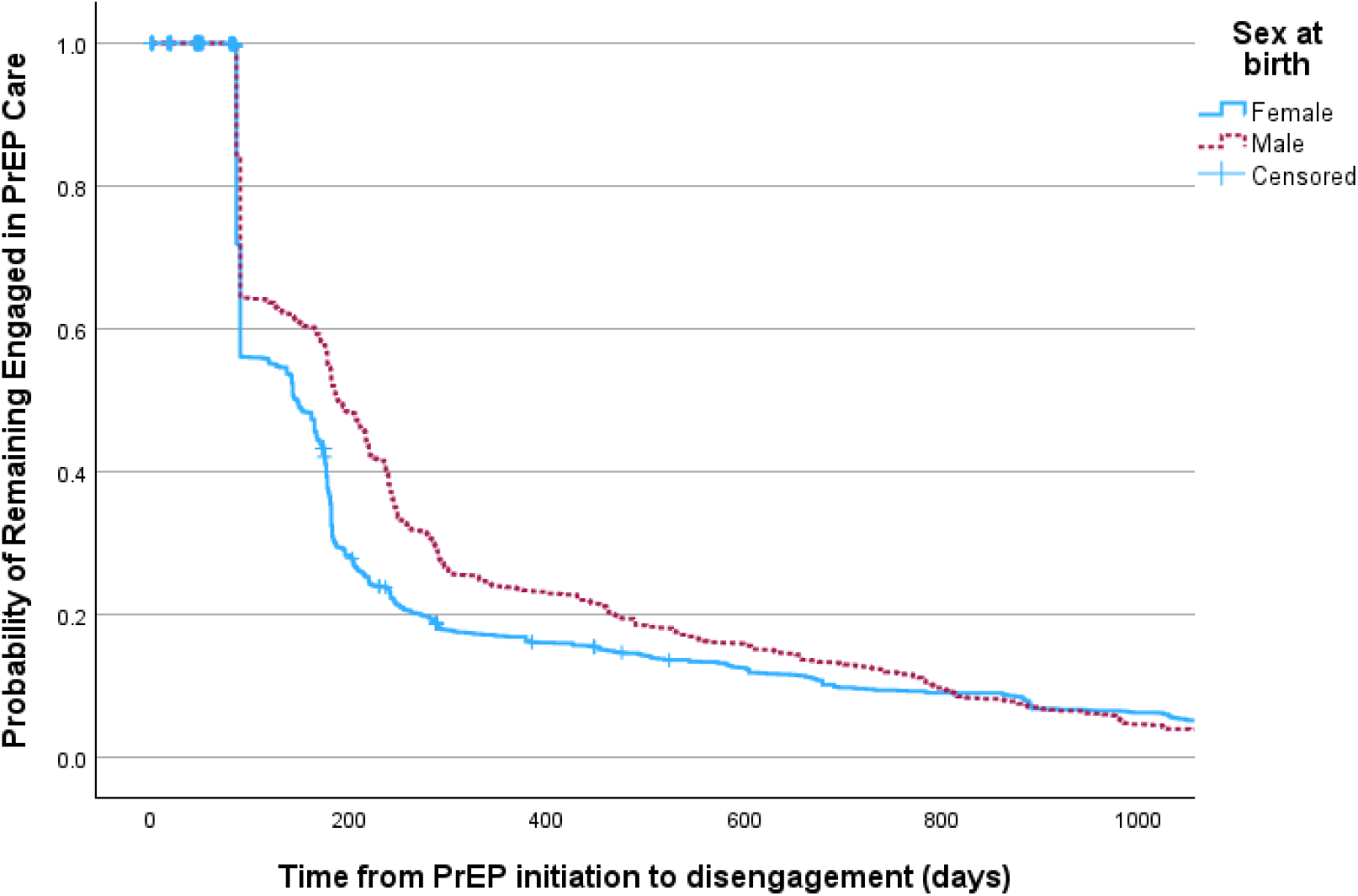
Kaplan–Meier survival curves for time to PrEP disengagement by sex at birth. Survival distributions also differed significantly by sex at birth (log-rank χ² = 35.86, df = 1, p < 0.001)

**Figure 2d.**
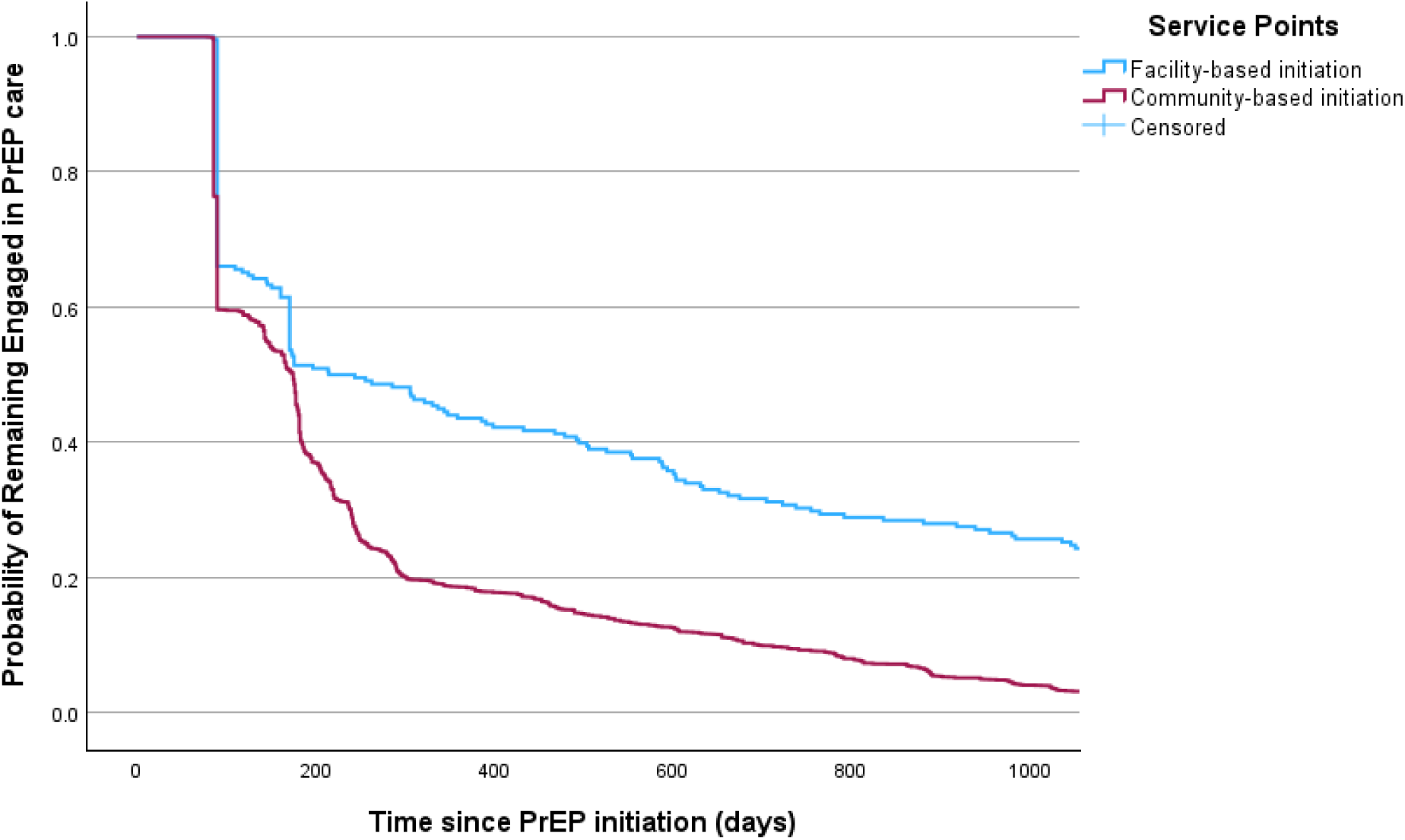
Kaplan–Meier survival curves for time to PrEP disengagement by service points.

### Factors associated with time to PrEP disengagement (Unadjusted Cox Models)

In bivariable Cox proportional hazards analyses, several individual, interpersonal, and service level factors were significantly associated with time to PrEP disengagement (Table 3). Participants who reported knowing their partner’s HIV status had a higher hazard of disengagement compared with those who did not know their partner’s status (HR = 1.80; 95% CI: 1.62–1.99; *p* < 0.001), and initiation through community-based service delivery points was also associated with a higher hazard relative to facility-based services (HR = 2.26; 95% CI: 1.94–2.63; *p* < 0.001). Significant differences were observed across population categories, with MSM (HR = 0.73; 95% CI: 0.67–0.78), PWID (HR = 0.71; 95% CI: 0.62– 0.80), and priority populations (HR = 0.45; 95% CI: 0.39–0.51) experiencing lower hazards of disengagement compared with sex workers (all *p* < 0.001). Peer outreach (follow-up) contact was strongly protective, with participants contacted by peer outreach workers exhibiting a substantially lower hazard of disengagement (HR = 0.31; 95% CI: 0.28–0.35; *p* < 0.001). Additional protective associations were observed among participants with any STI syndrome at baseline (HR = 0.43; 95% CI: 0.33–0.55; *p* < 0.001), those reporting psychoactive drug use in the prior six months (HR = 0.86; 95% CI: 0.76–0.98; *p* = 0.023), and those reporting experiences of gender-based violence (HR = 0.80; 95% CI: 0.65–1.00; *p* = 0.050). Sociodemographic factors showed more modest associations: older young adults aged 25–29 years had a slightly higher hazard of disengagement compared with adolescents aged 15–19 years (HR = 1.11; 95% CI: 1.00–1.22; *p* = 0.043), secondary education was associated with a higher hazard relative to no or primary education (HR = 1.16; 95% CI: 1.08–1.25; *p* < 0.001), Muslim participants had a lower hazard than Christians (HR = 0.85; 95% CI: 0.79–0.92; *p* < 0.001), and marital status was associated with disengagement, with married participants showing lower risk (HR = 0.78; 95% CI: 0.73–0.85; *p* < 0.001) and previously married participants higher risk (HR = 1.16; 95% CI: 1.05–1.28; *p* = 0.003). Sex at birth was not significantly associated with time to disengagement.

**Table 3.** Unadjusted Cox proportional hazards models for time to PrEP disengagement (N = 3,553)

| Variable | uHR (95% CI) | p-value |
| --- | --- | --- |
| **Sex at birth** |  |  |
| Female (ref) | 1 | — |
| Male | 1.02 (0.92–1.14) | 0.691 |
| **Education level** |  |  |
| None / Primary (ref) | 1 | — |
| Secondary | 1.16 (1.08–1.25) | <0.001 |
| Tertiary | 1.07 (0.93–1.25) | 0.354 |
| **Religion** |  |  |
| Christian (ref) | 1 | — |
| Muslim | 0.85 (0.79–0.92) | <0.001 |
| **Marital status** |  |  |
| Single (ref) | 1 | — |
| Married | 0.78 (0.73–0.85) | <0.001 |
| Previously married† | 1.16 (1.05–1.28) | 0.003 |
| **Population category** |  |  |
| Sex workers (ref) | 1 | — |
| Men who have sex with men (MSM) | 0.73 (0.67–0.78) | <0.001 |
| People who inject drugs (PWID) | 0.71 (0.62–0.80) | <0.001 |
| Priority populations | 0.45 (0.39–0.51) | <0.001 |
| **Age group** |  |  |
| Adolescents (15–19 years) (ref) | 1 | — |
| Young adults (20–24 years) | 1.12 (1.01–1.24) | 0.035 |
| Older young adults (25–29 years) | 1.17 (1.05–1.31) | 0.004 |
| **Knows partner's HIV status** |  |  |
| Does not know (ref) | 1 | — |
| Knows partner's HIV status | 1.80 (1.62–1.99) | <0.001 |
| **Peer follow-up after initiation** |  |  |
| No (ref) | 1 | — |
| Yes | 0.80 (0.65–1.00) | <0.001 |
| **Any gender-based violence (GBV)** |  |  |
| No (ref) | 1 | — |
| Yes | 0.80 (0.65–1.00) | 0.05 |
| **Any STI syndrome at baseline‡** |  |  |
| No (ref) | 1 | — |
| Yes | 0.42 (0.32–0.55) | <0.001 |
| **Any psychoactive drug use (last 6 months)** |  |  |
| No (ref) | 1 | — |
| Yes | 0.79 (0.54–1.14) | 0.208 |
| **Service delivery point** |  |  |
| Facility-based (ref) | 1 | — |
| Community-based | 2.26 (1.94–2.63) | <0.001 |

HR = hazard ratio; CI = confidence interval; ref = reference category
† Previously married includes individuals who were divorced, separated, or widowed at baseline.
‡ Any STI syndrome at baseline indicates documentation of one or more sexually transmitted infection syndromes at the baseline visit (yes vs no).

### Factors associated with time to PrEP disengagement (Adjusted Cox Model)

In the multivariable Cox proportional hazards model adjusting for sociodemographic, behavioral, relational, and service delivery factors (Table 4; N = 3,391), several variables remained independently associated with time to PrEP disengagement. After adjustment, individuals who reported knowing their partner’s HIV status had a significantly higher hazard of disengagement compared with those who did not (adjusted hazard ratio [aHR] = 2.04; 95% CI: 1.82–2.29; p < 0.001). Service delivery modality was also independently associated with disengagement, with clients initiating PrEP in the community experiencing a higher hazard of disengagement compared with facility-based services (aHR = 1.42; 95% CI: 1.17–1.72; p < 0.001). Significant differences persisted across population categories, with people who inject drugs exhibiting a lower hazard of disengagement compared with sex workers (aHR = 0.75; 95% CI: 0.62–0.89; p = 0.001), while no statistically significant differences were observed for MSM after adjustment. Age group was independently associated with disengagement, with both young adults aged 20–24 years (aHR = 1.12; 95% CI: 1.01–1.24; p = 0.035) and older young adults aged 25–29 years (aHR = 1.17; 95% CI: 1.05–1.31; p = 0.004) exhibiting higher hazards of disengagement compared with adolescents aged 15–19 years. Marital status remained strongly associated with disengagement, with married participants experiencing a lower hazard compared with single participants (aHR = 0.69; 95% CI: 0.64–0.76; p < 0.001). In contrast, sex at birth, education level, and recent psychoactive drug use were not significantly associated with time to disengagement in the adjusted model. Reporting any STI syndrome at baseline was associated with a substantially lower hazard of disengagement (aHR = 0.42; 95% CI: 0.32–0.55; p < 0.001), as was reporting recent experience of gender-based violence (aHR = 0.76; 95% CI: 0.60–0.96; p = 0.023).

**Table 4.**
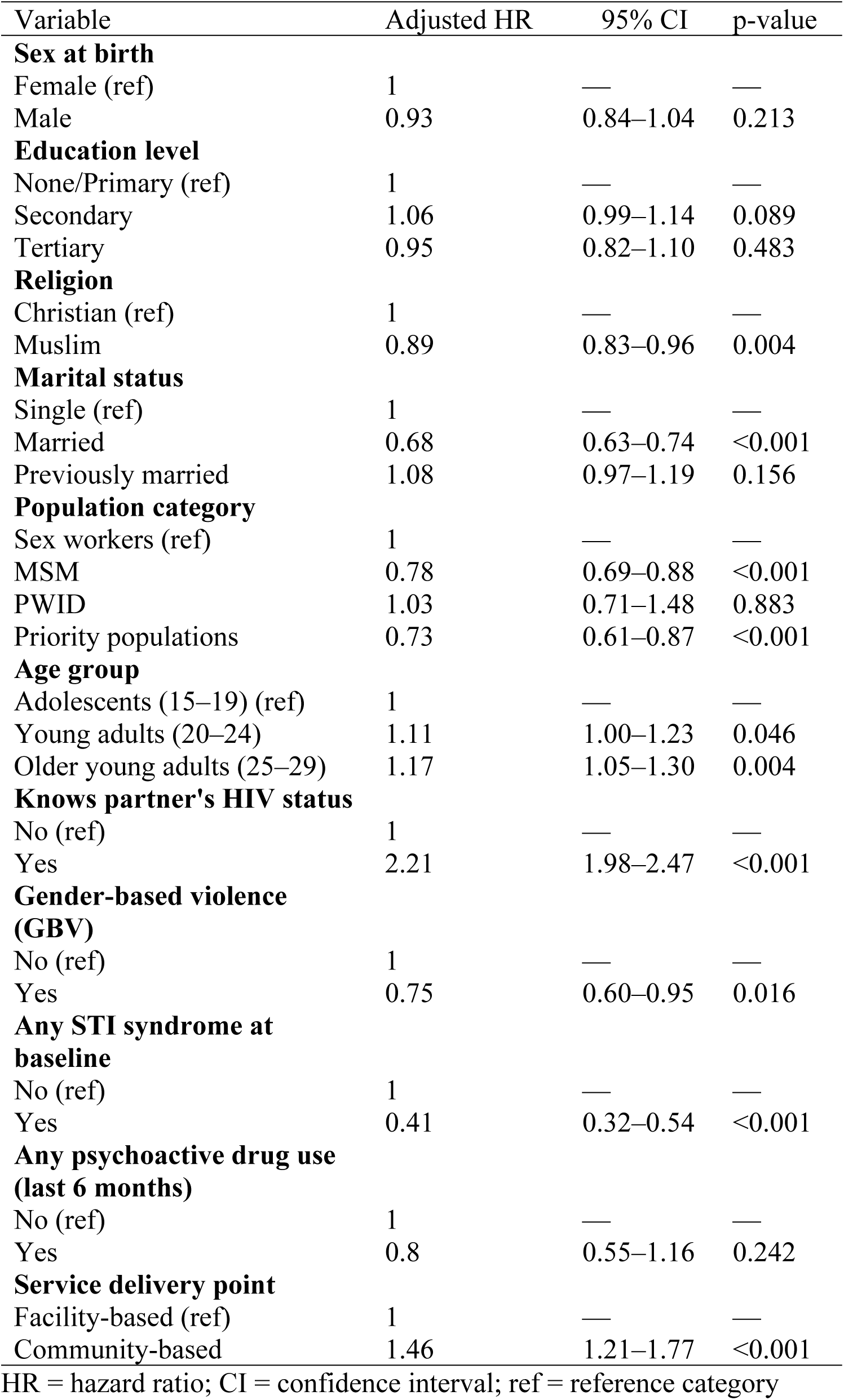
Factors Associated with Time to PrEP Disengagement: Multivariable Adjusted Cox Proportional Hazards Model (N = 3,391)

### Sensitivity analysis

To assess the robustness of the final adjusted Cox proportional hazards model, we conducted a theory-informed sensitivity analysis redefining the time origin to day 91 following PrEP initiation to reflect the programmatic 90-day grace period (Supplementary Table S1; N = 3,538). Findings remained highly consistent with the primary adjusted Cox model. Knowledge of a partner’s HIV status continued to be strongly associated with faster PrEP disengagement (aHR = 2.21; 95% CI: 1.98–2.47; *p* < 0.001), as did initiation through community-based service delivery compared with facility-based services (aHR = 1.46; 95% CI: 1.21–1.77; *p* < 0.001). Marital status remained a significant predictor, with married participants exhibiting a lower hazard of disengagement compared with single participants (aHR = 0.68; 95% CI: 0.63–0.74; *p* < 0.001), while no significant association was observed for previously married individuals. Differences by population category persisted, with men who have sex with men (aHR = 0.78; 95% CI: 0.69–0.88; *p* < 0.001) and priority populations (aHR = 0.73; 95% CI: 0.61–0.87; *p* < 0.001) demonstrating lower hazards of disengagement compared with sex workers, whereas no significant difference was observed for PWID. Age-related patterns were also consistent, with both young adults aged 20–24 years (aHR = 1.11; 95% CI: 1.00–1.23; *p* = 0.046) and older young adults aged 25–29 years (aHR = 1.17; 95% CI: 1.05–1.30; *p* = 0.004) exhibiting higher hazards of disengagement compared with adolescents aged 15–19 years. STI syndrome at baseline remained associated with substantially lower disengagement (aHR = 0.41; 95% CI: 0.32–0.54; *p* < 0.001), as did reporting recent gender-based violence (aHR = 0.75; 95% CI: 0.60–0.95; *p* = 0.016). In contrast, sex at birth, education level, and recent psychoactive drug use were not significantly associated with time to PrEP disengagement in the sensitivity model. Overall, the consistency of effect estimates across the primary and sensitivity analyses indicates that the observed associations were robust to alternative specifications of the risk period.

## Discussion

Among adolescents and young adults from key and priority populations accessing HIV prevention services in eastern Uganda, this study demonstrates substantial early disengagement from oral pre-exposure prophylaxis under routine program conditions. Although PrEP initiation was high among eligible clients, engagement declined rapidly, with only about one in five individuals remaining engaged at 12 months, underscoring the challenge of sustaining PrEP use in a population with elevated and fluctuating HIV risk. Similar patterns of high uptake followed by rapid disengagement have been reported across PrEP programs in sub-Saharan Africa have been reported in Uganda and beyond. For example, Kagaayi et al. (2020) documented early discontinuation among key and priority populations during PrEP rollout, with a median time on PrEP of approximately 45 days, indicating substantial drop-off within the first two months of initiation while Kawuma et al. (2025) observed marked declines in continuation over time, with fewer than half of participants retained in care by six months following PrEP initiation. Together, this evidence suggests that PrEP initiation among adolescents and young adults from key populations often occurs during periods of heightened concern. PrEP use is therefore conceptualized as situational rather than continuous, particularly in the context of changing partnerships, mobility, STI infection, and competing social and economic priorities. While such risk-responsive use may be appropriate in some circumstances, the steep decline in engagement observed in this study highlights a mismatch between current PrEP delivery models and the lived risk trajectories of adolescents and young adults from key populations. This mismatch may be reinforced by programmatic implementation models that prioritize PrEP initiation targets over sustained engagement, potentially limiting the time and support available to prepare clients for continued use. These findings underscore the need for prevention approaches that move beyond a narrow focus on initiation and instead support sustained, flexible, and risk-aligned PrEP use over time through counseling that addresses changing HIV risk, clear pathways for discontinuation and re-initiation, and differentiated service delivery models that reduce barriers to continued engagement. Initiation of PrEP through community-based service delivery points was associated with earlier disengagement compared with initiation at facility-based services, underscoring the role of service delivery modality in shaping continuity of PrEP use. Community-based models are highly effective for expanding access and facilitating initiation among adolescents and young adults from key and priority populations by reaching individuals in convenient, risk-responsive settings (Bogart et al., 2024). However, community initiation often occurs through mobile, outreach, or hotspot-based platforms that prioritize rapid enrollment, which may limit opportunities for sustained follow-up, ongoing risk counseling, and continuity of care. Structural barriers common to community settings, including high client mobility, reliance on episodic contact points, weaker tracking systems, and financing models that prioritize initiation outputs over retention may inadvertently constrain long-term engagement, particularly when follow-up activities rely on external funding (Haberer et al., 2017; Koss et al., 2021). In contrast, facility-based services may offer more structured follow-up, stronger patient–provider relationships, and better integration with routine clinical monitoring, which can support continued PrEP use among individuals with ongoing risk. However, these services often operate within overstretched health systems that limit privacy and provider time (Celum & Baeten, 2020; UNAIDS, 2021). Together, these findings highlight the importance of distinguishing between platforms optimized for PrEP initiation and those better suited for sustaining use. Community-based PrEP delivery should therefore be paired with explicit retention strategies, including hybrid community–facility models, peer- or digital-supported follow-up, flexible refill pathways, and strengthened linkage mechanisms. More broadly, differentiated service delivery approaches should be designed and evaluated with persistence rather than initiation alone as a core programmatic outcome for adolescents and young adults from key populations.

Retention patterns differed markedly across population groups, revealing important structural inequities in PrEP persistence. Compared with sex workers, men who have sex with men and other priority populations demonstrated more sustained engagement, while people who inject drugs showed no consistent differences after accounting for other factors. In contrast, sex workers experienced consistently earlier disengagement, underscoring the disproportionate challenges this group faces in maintaining PrEP use over time. These findings suggest that disengagement is driven less by individual adherence behavior and more by the broader risk environments in which PrEP is used. Sex workers often navigate high mobility, criminalization, stigma, exposure to violence, and competing economic and survival priorities, all of which can undermine continuity of prevention care despite ongoing HIV risk (UNAIDS, 2024). The observed disparities highlight limitations of uniform PrEP delivery models that assume similar capacity for sustained engagement across populations. Instead, they point to the need for equitable, differentiated, human-centered, responsive persistence strategies that reduce the burden of continued PrEP use in structurally vulnerable settings. Approaches such as peer-led support, flexible refill, and peer-to-peer follow-up mechanisms, while not fully captured in the final analytic model, may be particularly important for supporting persistence among sex workers and other highly marginalized groups. From an equity perspective, these retention gaps reflect unequal structural conditions shaping HIV risk and prevention access, reinforcing the need for PrEP persistence interventions that are explicitly tailored to population-specific realities rather than uniformly applied across diverse risk contexts.

Partner HIV testing was associated with a greater likelihood of PrEP discontinuation, suggesting that knowledge of a partner’s HIV status may prompt re-evaluation of perceived HIV risk rather than sustained prevention engagement. When uncertainty about a partner’s status is resolved, such as confirmation of HIV-negative results or awareness that a partner living with HIV is stable on ART treatment with undetectable viral load, individuals may interpret their immediate risk as reduced and subsequently disengage from PrEP use. This pattern challenges the assumption that partner testing consistently reinforces PrEP persistence and instead highlights the fluid and context-dependent nature of HIV risk perception among young KPs (Mugwanya et al., 2016; Davey et al., 2020). For young people engaged in changing or non-exclusive partnerships, PrEP may be understood as a temporary response to periods of heightened concern rather than as ongoing protection against future exposure. These dynamics are particularly important for sex workers, for whom reliance on a single partner’s HIV test result may provide an incomplete assessment of risk and inadvertently contribute to premature PrEP discontinuation despite continued exposure across multiple partnerships. In the Ugandan setting, routine HIV testing during PrEP use is intended to confirm continued eligibility or facilitate timely transition to HIV treatment if infection occurs, while partner testing is primarily used to inform prevention counseling and risk assessment rather than to signal risk elimination (MoH, 2018; MoH, 2022; WHO, 2016). Taken together, these findings underscore the importance of counseling approaches that clearly distinguish risk assessment from risk resolution and reinforce PrEP as an individual-controlled strategy for ongoing and future protection. For sex workers and other key populations, this may include complementary prevention approaches such as broader partner notification strategies, index and partner self-testing, and consistent condom use to address cumulative risk across partnerships (Okumu et al., 2022), while recognizing that some individuals may conceptualize PrEP use as episodic during periods of heightened vulnerability. Strategies to promote PrEP persistence may therefore need to be tailored to the specific needs and risk contexts of adolescents and young adults from key populations, ensuring that partner testing supports informed decision-making without inadvertently encouraging disengagement from PrEP in the presence of ongoing or evolving HIV risk (UNAIDS, 2021).

Reporting STI syndromes or recent exposure to gender-based violence at baseline was associated with greater PrEP persistence, revealing counterintuitive but important protective patterns. Rather than indicating vulnerability that undermines engagement, these markers may reflect heightened risk awareness, increased clinical contact, and more intensive provider follow-up, all of which can strengthen continuity of PrEP use. Individuals presenting with STIs or GBV are more likely to receive targeted counseling, referrals, and closer monitoring, potentially reinforcing the perceived relevance of PrEP and facilitating sustained engagement over time (World Health Organization, 2016; UNAIDS, 2021). Similar patterns have been observed in prior implementation studies where individuals with documented risk markers exhibited greater motivation to persist on PrEP (Ware et al., 2019; Rousseau et al., 2021). These findings suggest that clinical and psychosocial contact points, often framed solely as indicators of elevated HIV risk, may also function as anchors for prevention continuity. They highlight opportunities to more intentionally integrate PrEP persistence support within STI and GBV service platforms, including enhanced counseling, linkage to supportive services, and coordinated follow-up. From a syndemic and trauma-informed perspective, addressing intersecting sexual health and violence-related needs alongside HIV prevention may reduce disengagement by aligning PrEP delivery with the lived realities of individuals facing compounded vulnerabilities (Logie et al., 2020). Collectively, these findings underscore the potential for STI and GBV services to serve not only as entry points for PrEP initiation but also as critical platforms for sustaining engagement among adolescents and young adults from key populations.

## Strengths and limitations

This study draws on a large, routinely collected programmatic dataset, enabling examination of PrEP persistence under real-world delivery conditions and enhancing the relevance of policy and implementation. The extended five-year observation period supports robust time-to-disengagement analyses and justifies an analytic focus on observed engagement patterns rather than inconsistently measured, time-varying reasons for discontinuation. By focusing on PrEP disengagement rather than initiation, the study addresses a critical but underexamined stage of the PrEP cascade among adolescents and young adults from key and priority populations with elevated and dynamic HIV risk. At the same time, reliance on routine data may have resulted in incomplete or misclassified information, and disengagement from care may not fully capture PrEP discontinuation, particularly in a highly mobile population where some individuals may have continued PrEP at other facilities. Important time-varying drivers of persistence, including changes in risk context, mobility, and psychosocial factors, were not consistently measured; therefore, findings from this regional program may not be fully generalized to other settings. Despite these limitations, the study provides robust implementation-relevant evidence on structural and population-level drivers of PrEP disengagement that can inform more equitable and persistence-focused PrEP delivery strategies.

## Conclusion

This study demonstrates that while PrEP initiation among adolescents and young adults from key and priority populations can be achieved at scale under routine program conditions, sustaining engagement remains a major challenge. Rapid disengagement reflects dynamic risk perceptions, structural and financing constraints, and delivery models optimized for initiation rather than persistence, with pronounced inequities for sex workers and differential effects by service modality and population group. At the same time, clinical and psychosocial contact points, such as STI and GBV services, appear to strengthen retention, highlighting opportunities to anchor PrEP persistence within integrated, trauma-informed, and syndemic-aware care. Together, these findings underscore the need to reorient PrEP programs toward persistence as a core outcome, through differentiated, population-responsive strategies that align counseling, follow-up, and service delivery with fluctuating risk and mobility. Strengthening hybrid community–facility pathways, peer-supported follow-up, and flexible re-initiation models will be essential to translating high PrEP uptake into sustained HIV prevention impact for adolescents and young adults from key populations.

## Data Availability

The data used in this study are not publicly available because they contain potentially identifiable individual-level health information and are subject to ethical and institutional data protection requirements. Access to the de-identified dataset may be considered upon reasonable request and approval from the relevant institutional ethics committees and data custodians. The authors confirm that all procedures for data access and use comply with applicable ethical and regulatory requirements.

## Acknowledgement

We thank the Ministry of Health for granting permission to conduct this secondary analysis and the management and staff of Mbale Regional Referral Hospital for their collaboration and commitment to service delivery and data reporting.

---

**Supplementary Table S1.**
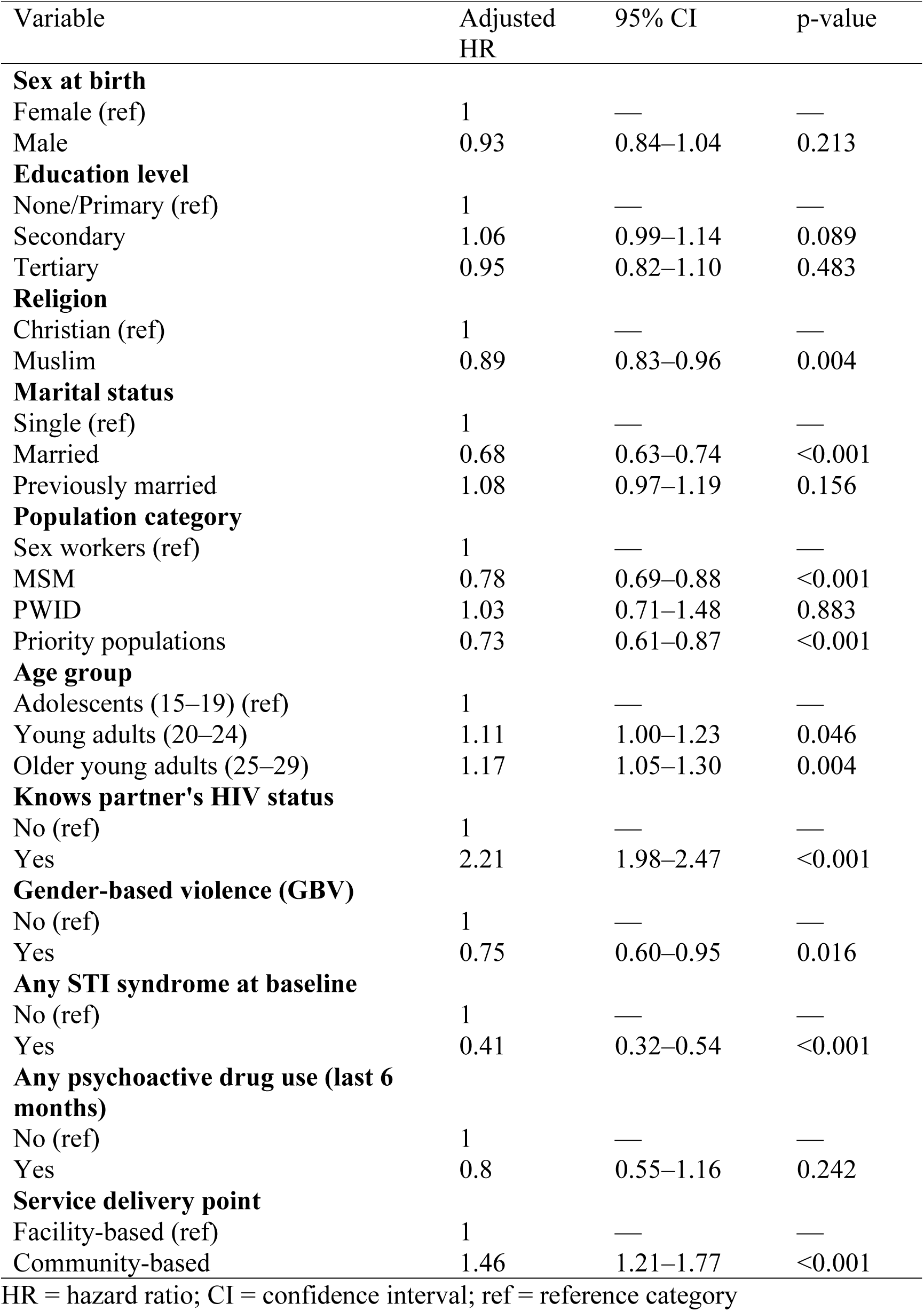
Sensitivity Analysis: Adjusted Cox Proportional Hazards Model for Time to PrEP Disengagement Using a 90-Day Time Origin (N = 3,538)

## References

1. Baeten, J. M., Heffron, R., Kidoguchi, L., Mugo, N. R., Katabira, E., Bukusi, E. A., Asiimwe, S., Haberer, J. E., Morton, J., Ngure, K., Bulya, N., Odoyo, J., Tindimwebwa, E., Hendrix, C., Marzinke, M. A., Ware, N. C., Wyatt, M. A., Morrison, S., Haugen, H., … Team, P. D. P. (2016). Integrated Delivery of Antiretroviral Treatment and Pre-exposure Prophylaxis to HIV-1–Serodiscordant Couples: A Prospective Implementation Study in Kenya and Uganda. PLOS Medicine, 13(8), e1002099. 10.1371/journal.pmed.1002099

2. Baral, S., Logie, C. H., Grosso, A., Wirtz, A. L., & Beyrer, C. (2013). Modified social ecological model: A tool to guide the assessment of the risks and risk contexts of HIV epidemics. BMC Public Health 2013 13:1, 13(1), 1–8. 10.1186/1471-2458-13-482

3. Bogart, L. M., Musoke, D. K., Allupo, S, Klein, D. J., Sejjemba, A., Mwima, S., Kadama, H., Mulebeke, R., & Wanyenze, R. K. (2024). Enhanced Oral Pre-exposure Prophylaxis (PrEP) Implementation for Ugandan Fisherfolk: Pilot Intervention Outcomes.

4. Corneli, A. L., Deese, J., Wang, M., Taylor, D., Ahmed, K., Agot, K., Lombaard, J., Manongi, R., Kapiga, S., Kashuba, A., & Van Damme, L. (2014). FEM-PrEP: adherence patterns and factors associated with adherence to a daily oral study product for pre-exposure prophylaxis. Journal of Acquired Immune Deficiency Syndromes (1999), *66*(3), 324–331. 10.1097/QAI.0000000000000158

5. Cowan, F. M., Davey, C., Fearon, E., Mushati, P., Dirawo, J., Chabata, S., Cambiano, V., Napierala, S., Hanisch, D., Wong-Gruenwald, R., Masuka, N., Mabugo, T., Hatzold, K., Mugurungi, O., Busza, J., Phillips, A., & Hargreaves, J. R. (2018). Targeted combination prevention to support female sex workers in Zimbabwe accessing and adhering to antiretrovirals for treatment and prevention of HIV (SAPPH-IRe): A cluster-randomised trial. The Lancet HIV, 5(8), e417–e426. 10.1016/S2352-3018(18)30111-5

6. Eakle, R., Gomez, G. B., Naicker, N., Bothma, R., Mbogua, J., Cabrera Escobar, M. A., Saayman, E., Moorhouse, M., Venter, W. D. F., Rees, H., & TAPS Demonstration Project Team. (2017). HIV pre-exposure prophylaxis and early antiretroviral treatment among female sex workers in South Africa: Results from a prospective observational demonstration project. PLoS Medicine, 14(11), e1002444. 10.1371/journal.pmed.1002444

7. Grant, R. M., Lama, J. R., Anderson, P. L., McMahan, V., Liu, A. Y., Vargas, L., Goicochea, P., Casap\’\ia, M., Guanira-Carranza, J. V., Ramirez-Cardich, M. E., & others. (2010). Preexposure chemoprophylaxis for HIV prevention in men who have sex with men. New England Journal of Medicine, 363(27), 2587–2599.

8. Heffron, R., Ngure, K., Mugo, N., Celum, C., Kurth, A., Curran, K., & Baeten, J. M. (2012). Willingness of Kenyan HIV-1 serodiscordant couples to use antiretroviral based HIV-1 prevention strategies. Journal of Acquired Immune Deficiency Syndromes (1999), *61*(1), 116–119. 10.1097/QAI.0b013e31825da73f

9. Kagaayi, J., Batte, J., Nakawooya, H., Kigozi, B., Nakigozi, G., Strömdahl, S., Ekström, A. M., Chang, L. W., Gray, R., Reynolds, S. J., Komaketch, P., Alamo, S., & Serwadda, D. (2020). Uptake and retention on HIV pre-exposure prophylaxis among key and priority populations in South-Central Uganda. Journal of the International AIDS Society, 23(8), e25588. 10.1002/jia2.25588

10. Kawuma, S., Katwesigye, R., Walusaga, H., Akatukunda, P., Nangendo, J., Kabugo, C., Kamya, M. R., & Semitala, F. C. (2025). Determinants of continuation on HIV pre-exposure propylaxis among female sex workers at a referral hospital in Uganda: A mixed methods study using COM-B model. BMC Public Health, 25(1), 143. 10.1186/s12889-024-20975-y

11. Kyongo, J. K. K., Karuga, R., Ochieng, C. N., Wachili, C., & Mukoma, W. (2018). How long will they take it? Oral pre-exposure prophylaxis (PrEP) retention for female sex workers, men who have sex with men and young women in a demonstration project in Kenya*. Vol.* 21, 54–55.

12. Lichtwarck, H. O., Mbotwa, C. H., Kazaura, M. R., Moen, K., & Mmbaga, E. J. (2023). Early disengagement from HIV pre-exposure prophylaxis services and associated factors among female sex workers in Dar es Salaam, Tanzania: A socioecological approach. BMJ Global Health, 8(12). 10.1136/bmjgh-2023-013662

13. Logie, C. H., Okumu, M., Mwima, S., Kyambadde, P., Hakiza, R., Kibathi, I. P., & Kironde, E. (2020). Sexually transmitted infection testing awareness, uptake and diagnosis among urban refugee and displaced youth living in informal settlements in Kampala, Uganda: A cross-sectional study. BMJ Sexual & Reproductive Health, 46(3), 192–199. 10.1136/bmjsrh-2019-200392

14. McCormack, S., Dunn, D. T., Desai, M., Dolling, D. I., Gafos, M., Gilson, R., Sullivan, A. K., Clarke, A., Reeves, I., Schembri, G., Mackie, N., Bowman, C., Lacey, C. J., Apea, V., Brady, M., Fox, J., Taylor, S., Antonucci, S., Khoo, S. H., … Gill, O. N. (2016). Pre-exposure prophylaxis to prevent the acquisition of HIV-1 infection (PROUD): Effectiveness results from the pilot phase of a pragmatic open-label randomised trial. The Lancet, 387(10013), 53–60. 10.1016/S0140-6736(15)00056-2

15. MoH. (2022a). *Consolidated guidelines for the prevention and treatment of HIV and AIDS in Uganda, November* 2022. https://files-hivpreventioncoalition.unaids.org/gpc/attachments/consolidated-guidelines-for-the-prevention-and-treatment-of-hiv-and-aids-in-uganda-2022.pdf

16. MoH. (2022b). Technical Guidance on PrEP for Persons at Substantial Risk of HIV in Uganda, 2022. PrEPWatch. https://www.prepwatch.org/resources/technical-guidance-on-prep-for-persons-at-substantial-risk-of-hiv-in-uganda-2022/

17. Mwima, S., Bogart, L. M., Musoke, W., Mukama, S. C., Allupo, S., Kadama, H., Naigino, R., Mukasa, B., & Wanyenze, R. K. (2025). Applying implementation science frameworks to understand why fisherfolk continue or discontinue pre-exposure prophylaxis for HIV prevention in Uganda: A qualitative analysis. BMJ Global Health, 10(1). 10.1136/bmjgh-2024-017368

18. Ntabadde, K., Kagaayi, J., Ssempijja, V., Feng, X., Kairania, R., Lubwama, J., Ssekubugu, R., Yeh, P. T., Ssekasanvu, J., Tobian, A. A. R., Kennedy, C. E., Mills, L. A., Alamo, S., Kreniske, P., Santelli, J., Nelson, L. J., Reynolds, S. J., Chang, L. W., Nakigozi, G., & Grabowski, M. K. (2024). Pre-exposure prophylaxis (PrEP) knowledge, use, and discontinuation among Lake Victoria fisherfolk in Uganda: A cross-sectional population-based study. medRxiv, 2024.03.29.24305076. 10.1101/2024.03.29.24305076

19. Nunn, A. S., Brinkley-Rubinstein, L., Oldenburg, C. E., Mayer, K. H., Mimiaga, M., Patel, R., & Chan, P. A. (2017). Defining the HIV pre-exposure prophylaxis care continuum. AIDS, 31(5), 731. 10.1097/QAD.0000000000001385

20. Okumu, M., Logie, C. H., Ansong, D., Mwima, S., Hakiza, R., & Newman, P. A. (2022). Exploring the Protective Value of Using Sexting for Condom Negotiation on Condom use Determinants and Practices Among Forcibly Displaced Adolescents in the Slums of Kampala, Uganda. AIDS and Behavior, 26(11), 3538–3550. 10.1007/s10461-022-03677-7

21. Rosen, J. G., Rao, A., Kibuuka, H., Mwesigwa, B., Mirembe, G., Musingye, E., Abdulrahman, A., Baral, S. D., Crowell, T. A., Akom, E. E., & Rucinski, K. B. (2025). Patterns and Predictors of HIV Pre-exposure Prophylaxis (PrEP) Continuity Among Young Women Who Sell Sex in Uganda: A Group-Based Trajectory Modeling Approach. AIDS and Behavior. 10.1007/s10461-025-04993-4

22. UNAIDS. (2025). Joint United Nations Programme on HIV/AIDS (UNAIDS). (2025). Global AIDS update 2025. UNAIDS. https://www.unaids.org/sites/default/files/2025-07/2025-global-aids-update-JC3153_en.pdf?utm_source=chatgpt.com

23. Zimmermann, H. M., Eekman, S. W., Achterbergh, R. C., Schim van der Loeff, M. F., Prins, M., de Vries, H. J., Hoornenborg, E., Davidovich, U., & Consortium (H-TEAM), O. behalf of the A. P. P. team in the H. T. E. Am. (2019). Motives for choosing, switching and stopping daily or event-driven pre-exposure prophylaxis – a qualitative analysis. Journal of the International AIDS Society, 22(10), e25389. 10.1002/jia2.25389

